# Emotional and behavioural pathways to adolescent substance use and antisocial behaviour: results from the UK Millennium Cohort Study_PREPRINT

**DOI:** 10.1101/2020.02.20.20024158

**Authors:** João Picoito, Constança Santos, Carla Nunes

## Abstract

This study examines the emotional and behavioural pathways to adolescent substance use and antisocial behaviour. Using a sample of 17,223 participants from the UK Millennium Cohort Study, we applied parallel-process growth mixture modelling on emotional and behavioural symptoms in those aged 3 to 14 and employed latent class analysis to identify patterns of substance use and antisocial behaviours at age 14. We then performed a multinomial regression analysis to explore the association between emotional and behavioural trajectories and patterns of adolescent substance use and antisocial behaviours, controlling for sociodemographic, family and maternal factors. We found five trajectories of emotional and behavioural symptoms and four classes of adolescence substance use and antisocial behaviour. Children and adolescents in the ‘*high externalising and internalising’* and *‘moderate externalising’* trajectories were more likely to belong to any problematic behaviour class, especially the ‘*poly-substance use and antisocial behaviours’* class. Inclusion in the *‘moderate externalising and internalising (childhood limited)’* class was associated with higher odds of belonging to the *‘alcohol and tobacco’* class. These associations remained significant after controlling for important sociodemographic and contextual factors, such as maternal substance use, poverty, and parental status. Interventions on adolescent health promotion and risk behaviour prevention need to address the clustering of substance use and antisocial behaviour as well as the significant influence of early and chronic internalising and externalising symptoms on the aetiology of these behaviours.

## Introduction

Adolescent substance use has a profound impact on both the individual and societal levels, as it is associated with negative health consequences and personal functional outcomes [1]. Substance use that starts early in life is associated with a greater risk of substance abuse and dependence later in life [1]. Other risk behaviours in adolescence, such as delinquency, antisocial behaviours, truancy and risky sexual behaviours, tend to cluster with substance use [2]. According to Jessor’s problem behaviour theory, this co-occurrence is explained by shared individual, psychological, family and cultural factors [3]. The interplay between these shared environmental factors triggers a developmental cascade that culminates in risk behaviours and delinquency in adolescence [4].

Risk factors and developmental pathways contributing to adolescent risk behaviours can be traced to early childhood. One line of research looks at the emotional and behavioural antecedents of adolescent substance use and antisocial behaviours [5]. Emotional and behavioural symptoms can be divided into externalising symptoms (conduct problems, hyperactivity, impulsivity and inattention) and internalising symptoms (depression, anxiety). Externalising symptoms in childhood and adolescence have been consistently associated with antisocial behaviours and substance use in adolescence [6–8]. The evidence for internalising symptoms is somewhat contradictory, however; some studies report positive associations [6] while others report negative findings [9]. Due to the dynamic nature of emotional and behavioural symptoms, the precise interplay between internalising and externalising symptoms from early childhood to adolescence and its associations with adolescent substance use and antisocial behaviour remains elusive.

The main objectives of this study are as follows: 1) identify patterns of substance use and antisocial behaviours at age 14; 2) identify joint trajectories of emotional problems, conduct problems and hyperactivity/inattention from ages 3 to 14; and 3) study the association of the trajectories of emotional problems, conduct problems and hyperactivity/inattention with patterns of adolescent substance use and antisocial behaviours, controlling for sociodemographic, family and maternal factors. To achieve these objectives, we used person-centred statistical methods, such as growth mixture modelling and latent class analysis, on a large general population sample.

## Methods

### Participants

The data for this study was obtained from the Millennium Cohort Study (MCS; http://www.cls.ioe.ac.uk/mcs), a population-based longitudinal study conducted in the United Kingdom. The purpose of the MCS is to assess the social, economic and health conditions of children born in the new century [10]. A stratified random sampling approach was used to overrepresent areas of high child poverty and ethnic minorities. A total of 19,243 families were enrolled in the MCS during the initial two sweeps (at 9 months and 3 years). Subsequent sweeps included children aged 5, 7, 11, and 14 years, including 15,246, 13,857, 13,287, and 11,726 families, respectively. Ethical approval for each wave was given by the NHS Multi-Centre Ethics Committees. Parents gave informed consent prior to the interviews. Further details on design, attrition and ethics of MCS can be found elsewhere [10]. For this study, we included cohort members (singletons and first-born twins or triplets) with valid data on emotional and behavioural problems in at least one of sweeps 2 through 6 (n=17,215) or on at least one measure of adolescent substance use and antisocial behaviours in sweep 6 (n=11,206). In total, there were 17,223 participants. Participants excluded from the analysis were more likely to be male, ethnic minorities, poor, and from single-parent families (Table S1).

### Measures

#### Adolescent substance use and antisocial behaviours

We assessed lifetime alcohol use, tobacco and cannabis. Lifetime alcohol use was assessed by asking “Have you ever had an alcoholic drink? That is more than a few sips”. Cannabis lifetime use was evaluated by asking “Have you ever tried any of the following things?” and presenting a list including cannabis. The answer categories for these three lifetime substance use measures were yes and no. Lifetime smoking was assessed by asking adolescents which one of six statements from “I have never smoked cigarettes” to “I usually smoke more than six cigarettes a week” best described them. Those answer categories were dichotomised into never and at least once.

Antisocial behaviours included physical fighting and property crimes such as shoplifting, vandalism and graffiti. Physical fighting was measured by asking “In the last 12 months, have you pushed or shoved/hit/slapped/punched someone?”. Shoplifting was measured by asking “In the last 12 months, have you taken something from a shop without paying for it?”. Vandalism was assessed by asking “In the last 12 months, have you purposely damaged anything in a public place that didn’t belong to you, for example by burning, smashing or breaking things like cars, bus shelters and rubbish bins”. Graffiti was measured by asking “In the last 12 months, have you written things or spray-painted on a building, fence or train or anywhere else where you shouldn’t have?”. Response categories were yes and no for all of the antisocial behaviour measures.

#### Emotional and behavioural symptoms

Participants’ emotional and behavioural symptoms were measured using three subscales from the Strengths and Difficulties Questionnaire (SDQ) [11]: emotional problems, conduct problems and hyperactivity/inattention. Each subscale comprises five items individually scored from 0 to 2—not true, somewhat true, certainly true—with the total score ranging from 0 to 10; higher scores indicate more problems or difficulties. Each subscale score was treated as a continuous variable. We used suggested cut-points for the measurement of severity: 5 for emotional problems, 4 for conduct problems and 7 for hyperactivity/inattention [12]. The cut-points for age 3 were lower for emotional problems (4) and higher for conduct problems (5) [13]. The reliability of the SDQ subscales used was good, with ordinal alpha [14] ranging across sweeps from 0.76 to 0.82 for emotional symptoms, 0.77 to 0.85 for conduct problems and 0.78 to 0.86 for hyperactivity/inattention.

#### Sociodemographic, family and maternal covariates

Contextual variables included gender, ethnicity (white versus other), family poverty (income below 60% of the national median), single-parent status and maternal professional qualifications (high versus other). Maternal alcohol use was assessed by asking “Which of these best describes how often you usually drink alcohol?” with answers ranging from “never” to “every day”. Responses were dichotomised into “never” and “at least less than once a month”. Maternal smoking was measured by asking “Do you smoke tobacco products such as cigarettes, cigars or a pipe at all nowadays?”, with responses including “no, does not smoke”, cigarettes, roll-ups, cigars, pipe and other. Response categories were dichotomised into “non-smoker” and “smoker”. Maternal psychosocial distress was assessed with a shortened, nine-item, self-completed version of the Rutter Malaise Inventory (Cronbach’s alpha=0.75) [15]. Family and maternal variables were collected at 9 months of age.

### Data analysis

First, we performed latent class analysis (LCA) of problem behaviour at age 14. To select the best model, we sequentially run models with one to six classes. We chose the best model assessing fit indices, specifically the Bayesian information criterion (BIC), with lower values reflecting better fit; the ‘elbow method’ to find the last largest relative decrease in the fit indices [16]. We also used the Lo-Mendell-Rubin test which assesses if an additional class significantly improves model fit and entropy, ranged between 0 and 1, with values >0.80 indicating less classification error. Interpretability and parsimony were considered when choosing the optimal number of classes. We tested the validity of the local independence assumption, using the residual association approach [17]. More details on these fit indices can be found elsewhere [18].

Second, we used a parallel-process growth mixture model (parallel-process GMM) to gauge the trajectories of emotional problems, conduct problems and hyperactivity/inattention jointly from ages 3 to 14 [2]. GMM is a type of latent growth curve model that identifies discrete groups of individuals within a population based on their longitudinal change [19]. We fitted a series of latent basis models with one to six classes. Latent basis is a flexible GMM framework that allows for freely estimating factor loadings to find the growth shape that optimally explains longitudinal change [20]. The best model fit was chosen following the same procedures for model selection described above for the LCA.

Third, we performed a multinomial regression of the adolescent problem behaviour classes on the parallel-process GMM classes of emotional and behavioural symptoms trajectories, controlling for sociodemographic, family and maternal covariates. We performed a corrected three-step analysis (using Mplus command r3step), adjusting for the classification error of the adolescent risk behaviour classes and preventing the covariates from influencing latent class formation [21]. For the parallel-process GMM classes, we adopted a classic classify-analyse approach, extracting modal class assignment for each observation and treating the parallel-process GMM classes as observed independent variables. When entropy is 0.80 or greater, some authors consider the classification error to be negligible [19]. For this final part of the analysis, we included the 11,198 participants with valid data on adolescent risk behaviours (8 participants with data on adolescent substance use and antisocial behaviours were excluded for not having any emotional or behavioural measures).

Missing data for the emotional problems, conduct problems and hyperactivity/inattention subscales of the SDQ as well as the age 14 problem behaviours were addressed with full-information maximum likelihood (FIML) procedures incorporated in parallel-process GMM and LCA; they were assumed to be missing at random. We additionally multiply imputed by chained equations 50 datasets to handle missing covariates [22]. The model of multiple imputations included all covariates, parallel-process GMM classes and substance use and antisocial behaviour indicators. The analyses accounted for the stratified clustered sample design and attrition of the MCS. We used Mplus version 8.3 [23] and packages MlusAutomation [24] and dotwhisker [25] for R version 3.5.1.

## Results

### Characteristics of the sample

From early childhood to adolescence, there was an increase in the mean score of emotional problems but a decrease in the mean score of conduct problems and hyperactivity/inattention (Table 1). Lifetime alcohol use at age 14 (49.6%) was far more common than tobacco (14.5%) and cannabis use (4.7%). Finally, while involvement in shoplifting (3.4%), graffiti (2.7%) and vandalism (3.0%) in the last 12 months was uncommon, involvement in physical fighting was somewhat prevalent (30.2%).

**Table 1.** Weighted descriptive statistics (n= 17223)

|  | n (%) /<br>mean (standard error) | Missing<br>n (%) |
| --- | --- | --- |
| **Emotional problems** |  |  |
| Age 3 | 1.31 (0.17) | 2480 (14.4) |
| Age 5 | 1.35 (0.02) | 2497 (14.5) |
| Age 7 | 1.49 (0.02) | 3780 (21.9) |
| Age 11 | 1.82 (0.03) | 4428 (25.7) |
| Age 14 | 1.97 (0.03) | 5896 (34.2) |
| **Conduct problems** |  |  |
| Age 3 | 2.75 (0.03) | 2453 (14.2) |
| Age 5 | 1.46 (0.02) | 2478 (14.4) |
| Age 7 | 1.34 (0.02) | 3751 (21.8) |
| Age 11 | 1.34 (0.02) | 4425 (25.7) |
| Age 14 | 1.35 (0.02) | 5894 (34.2) |
| **Hyperactivity/inattention** |  |  |
| Age 3 | 3.86 (0.03) | 2591 (15.0) |
| Age 5 | 3.23 (0.03) | 2564 (14.9) |
| Age 7 | 3.30 (0.04) | 3801 (22.1) |
| Age 11 | 3.05 (0.04) | 4452 (25.8) |
| Age 14 | 2.90 (0.04) | 5901 (34.3) |
| **Adolescence risk behaviours** |  |  |
| Lifetime alcohol use | 49.6 (0.8) | 6040 (35.1) |
| Lifetime tobacco use | 14.5 (0.5) | 6079 (35.3) |
| Lifetime cannabis use | 4.7 (0.2) | 6046 (35.1) |
| Physical fighting | 30.2 (0.5) | 6040 (35.1) |
| Shoplifting | 3.4 (0.2) | 6040 (35.1) |
| Graffiti | 2.7 (0.2) | 6034 (35.0) |
| Vandalism | 3.0 (0.2) | 6039 (35.1) |
| **Sociodemographic, family and maternal variables** |  |  |
| Gender (male) | 51.1 (0.5) | - |
| Ethnicity (white) | 87.8 (1.1) | 100 (0.6) |
| Single-parent family | 13.1 (0.5) | 673 (3.9) |
| Family income poverty | 27.6 (0.9) | 726 (4.2) |
| Mother's high professional qualification (NVQ 4 or 5) | 34.6 (1.1) | 703 (4.1) |
| Maternal alcohol use (any) | 81.9 (0.8) | 688 (4.0) |
| Maternal tobacco use (any) | 27.9 (0.7) | 688 (4.0) |
| Maternal psychosocial distress (Rutter's Malaise Inventory) | 1.58 (0.05) | 1281 (7.4) |

Results are presented as n [unweighted frequency] (%) [weighted percentage] for categorical variables, and as mean (standard error) for continuous variables.

### Adolescent substance use and antisocial behaviour latent classes

A five-class model provided the lowest BIC (Table S2). Additionally, with the six-class model, the first non-significant p-value for the LMRT emerged, supporting the choice of the five-class model. However, applying the principle of parsimony, and based on the good interpretability of the latent classes, we ultimately chose the four-class model as the optimal solution. Additionally, the last largest relative decrease in the fit indices was achieved with the four-class model.

Figure 1 shows the resulting four classes. The ‘*normative’* class, making up 71.9% of the sample, displayed a low probability of substance use and antisocial behaviours. ‘*Alcohol and physical fighting’* was the second most common class (15.2%) with a high probability of alcohol use and involvement in physical fights. The ‘*alcohol and tobacco’* class had the highest odds of a positive lifetime tobacco smoking and represented 9.9% of the sample. ‘*Poly-substance use and antisocial behaviours’* was the least common class (3%) with the highest probability of cannabis use and any antisocial behaviour.

**Figure 1.**
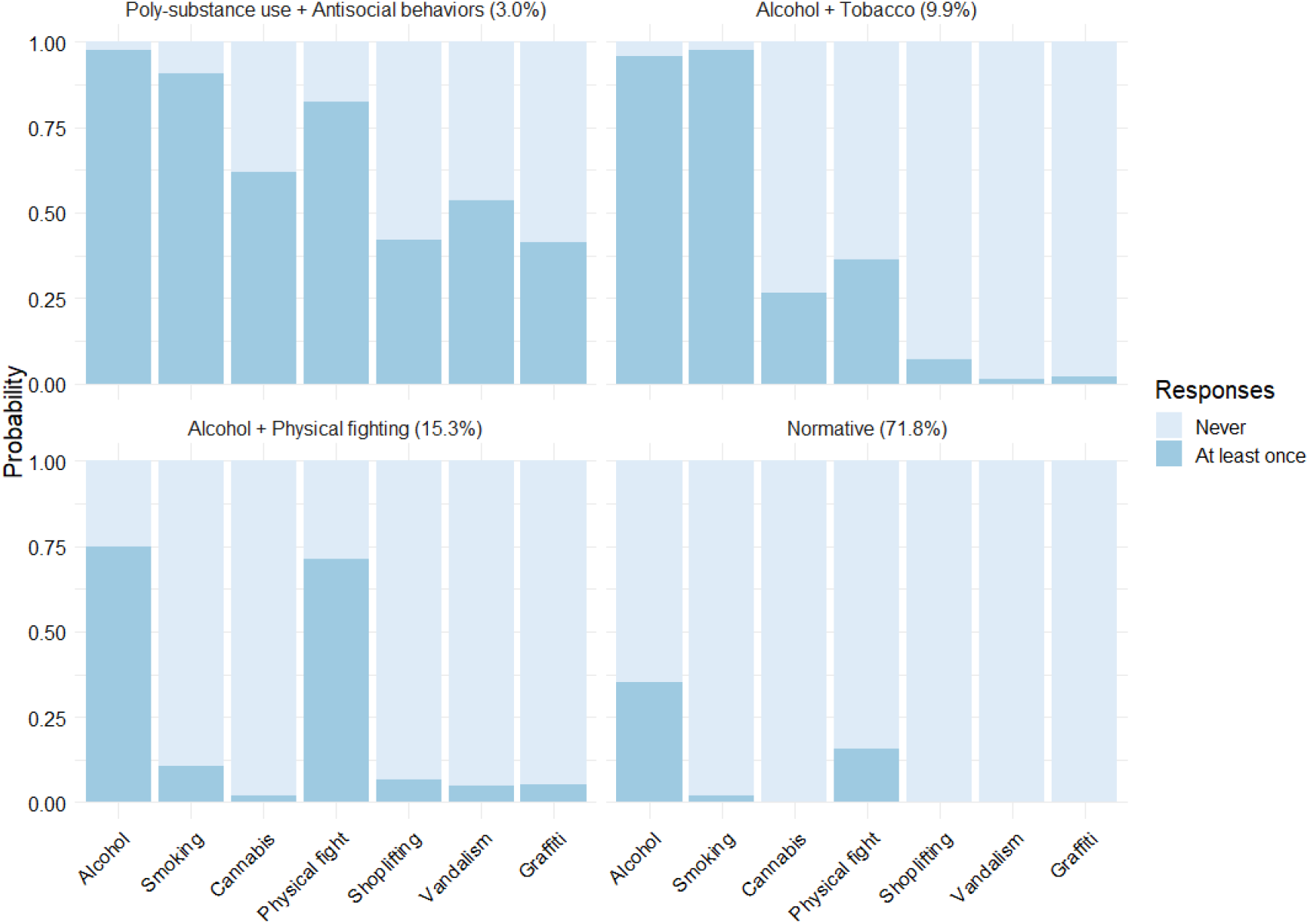
Substance use and antisocial behaviors profiles

### Emotional and behavioural symptoms trajectories

We estimated a series of parallel-process GMM models including 17,215 children with an increasing number of classes (Table S3). The fit indices did not reach a minimum value. The ‘elbow method’ suggested the five-class model as optimal. The interpretability of the resulting classes, good entropy (0.826), and significant LMRT p-value supported this suggestion.

Figure 2 shows the resulting five parallel-process GMM classes: (a) ‘*low symptoms’*, making up 73.5% of the children; (b) ‘*moderate externalising’*, with low emotional problems but borderline hyperactivity/inattention and conduct problems (8.9%); (c) ‘*moderate externalising and internalising (childhood limited)’*, similar to the previous class but with lower scores for emotional problems, conduct problems and hyperactivity/inattention (7.1%); (d) ‘*high internalising (preadolescence onset)’*, with an increasing emotional problems score peaking in adolescence (8.0%); and (e) ‘*high externalising and internalising’*, with the highest scores of emotional problems, conduct problems and hyperactivity/inattention throughout development (2.6%). Table S4 reports the mean intercept and slope for each trajectory.

**Figure 2.**
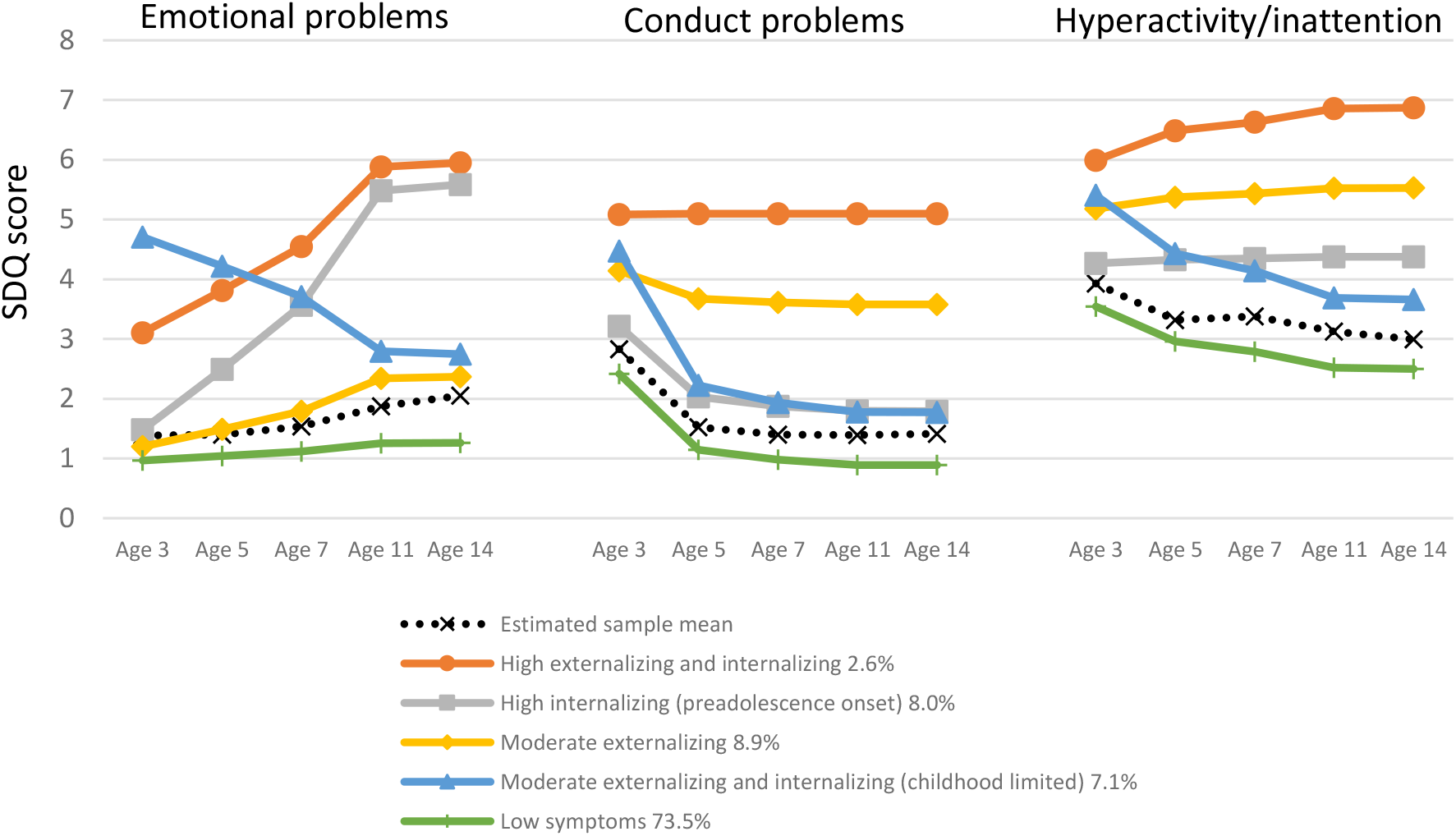
Emotional and behavioural symptoms trajectories. SDQ – Strengths and Difficulties Questionnaire.

### Associations between emotional and behavioural trajectories and adolescent problem behaviour latent classes

For this part of the analysis, we included a group of participants with valid data on adolescent problem behaviours and emotional and behavioural trajectories, comprising 11,198 of the initial 17,223 children. Participants excluded were more likely to be male, be from single-parent families and report lower levels of maternal professional qualifications. Excluded children also had higher levels of emotional problems, conduct problems and hyperactivity/inattention at all ages.

Figure 3 shows the associations between adolescent substance use latent classes and emotional and behavioural trajectories, controlling for a set of sociodemographic, family and maternal factors (for the unadjusted odds ratios, see figure S1). Children and adolescents in the ‘*high externalising and internalising’* and *‘moderate externalising’* classes were more likely to belong to a problem behaviour class, especially the ‘*poly-substance use and antisocial behaviours’* class, than to the ‘*normative’* class (OR 5.11, CI 2.61-9.99; OR 4.83, CI 3.17-7.35, respectively). Membership in the *‘moderate externalising and internalising (childhood limited)’* class was associated with higher odds of belonging to the *‘alcohol and tobacco’* class (OR 1.50, CI 1.05-2.14). Male gender was associated with higher odds of membership in the *‘poly-substance use and antisocial behaviours’* class (OR 1.99, CI 1.46-2.72) and the *‘alcohol and physical fighting’* class (OR 4.56, CI 3.40-6.10). Maternal alcohol use was related to all problem behaviour classes, especially *‘alcohol and physical fighting’* (OR 2.67, CI 1.68-4.24). Similarly, maternal tobacco use was associated with all problem behaviour classes but with higher odds for the *‘alcohol and tobacco’* (OR 2.85, CI 2.23-3.64) and *‘poly-substance use and antisocial behaviours’* classes (OR 2.49, CI 1.74-3.59). Maternal psychosocial distress was associated with higher odds for the *‘alcohol and tobacco’* class (OR 1.09, CI 1.02-1.16).

**Figure 3.**
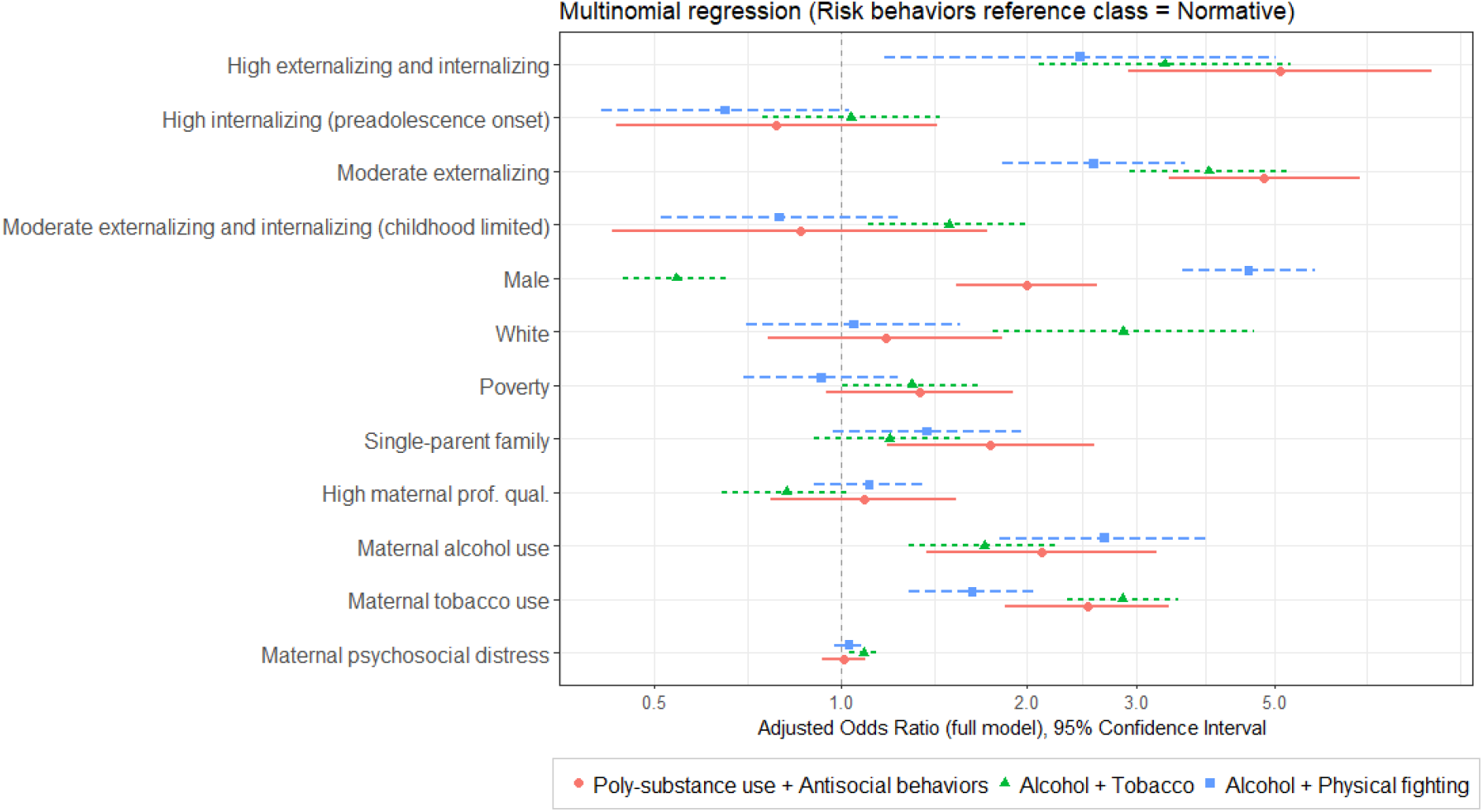
Associations between emotional and behavioural trajectories, sociodemographic, family and maternal factors (independent variables), and adolescence problem behaviour latent classes (outcome variables). Trajectories of emotional and behavioural symptoms reference class = Low symptoms. White (ref=other ethnicity); Poverty (ref=no family poverty); Single-parent (ref=other family types); high maternal professional qualification (ref=other maternal professional qualifications); Maternal alcohol use (ref=no maternal alcohol use); Maternal tobacco use (ref=no maternal tobacco use).

## Discussion

This study looked at the emotional and behavioural pathways to adolescent substance use and antisocial behaviour using a large general population sample. We found four patterns of adolescent problem behaviours; these ranged from the normative class with low substance use and antisocial behaviour to the poly-substance use class with high alcohol, tobacco, and cannabis use and high involvement in physical fighting and property crime offenses. There were two additional classes with high alcohol use and physical fighting as well as a class with high alcohol and tobacco use. We found five trajectories of emotional and behavioural symptoms; these included two with persistent symptoms from early childhood to adolescence, one with both high internalising and externalising symptoms and another with moderate externalising symptoms. There were also two patterns with transient symptoms, the childhood-limited moderate externalizing trajectory and the internalizing trajectory, as well as one with high internalizing symptoms with onset prior to adolescence. Children with persistent moderate or high externalising symptoms were more likely to engage in substance use and antisocial behaviours in adolescence relative to children with low emotional and behavioural symptoms. Even when the internalising and externalising symptoms were limited to childhood, these children had increased odds of alcohol and tobacco use in adolescence. These associations remained significant after controlling for important sociodemographic and contextual factors, such as maternal substance use, poverty and single-parent status.

We found two patterns of concomitant substance use and antisocial behaviours, namely ‘*alcohol and physical fighting’* and ‘*poly-substance use and antisocial behaviours’*. There are several reports in the literature on the close relationship between adolescent substance use and antisocial behaviours, both cross-sectionally [26–28] and longitudinally [29–32]. A study on substance use and delinquency clustering from adolescence to early adulthood identified a pattern of concurrent delinquency and substance use, in addition to a low risk and primary delinquent classes [31]. Using data from the 1970 British Cohort Study, Akasaki et al. [27] found a risky behaviour class in 16-year-old adolescents characterised by smoking, binge drinking and involvement in physical fighting. A previous study also found that adolescent risk behaviours, such as cannabis use, conduct problems, delinquency and early sexual activity, tended to cluster together, albeit with significant gender differences [26]. Using a parallel-process GMM, Wu et al. [2] found four trajectories of conduct problems and substance use, including a high conduct problem and high substance use class, and an increasing conduct problem and substance use class.

A recent study using data from the MCS analysed the longitudinal change in internalising and externalising symptoms as well as general cognitive ability from early childhood to preadolescence [33]. The authors found four classes: a typical developing class, a class with improving behaviour, a class with worsening behaviour, and a class with persistently high levels of internalising and externalising symptoms [33]. Our results further expand on these findings, parsing externalising behaviours into conduct problems and hyperactivity/inattention, allowing for the detection of more nuanced patterns of emotional and behavioural symptoms. High levels of conduct problems can start early in life and are associated with increasing levels of emotional problems—as well as increasing hyperactivity/inattention symptoms to a lesser degree— throughout development. This is in line with reports in the literature of a pattern of early-onset internalising and externalising symptoms that is associated with worse social and developmental outcomes later on [34–37]. In contrast to emotional and conduct problems, hyperactivity/inattention had a more stable course throughout development. A study on the developmental course of hyperactivity/inattention symptoms from early childhood to adolescence found a decrease in hyperactivity and impulsivity symptoms, while attention symptoms were consistent [38]. We did not, however, find the trajectory with peaking externalising symptoms in preadolescence or adolescence that some other studies found [34–36]. We instead discovered a pattern of emerging internalising problems in adolescence, which has also been reported [39, 40].

The association between externalising symptoms and adolescent risk behaviours has been consistently reported in the literature [41]. Externalising symptoms increase the risk of all substance use [6]. Conduct problems in childhood are strong predictors of delinquency and antisocial behaviour in adolescence and adulthood [7]. The timing of the emergence of externalising symptoms and conduct problems and its relationship with adolescence substance use has also been shown to indicate a higher risk for early-onset externalising behaviours [8]. In our study, trajectories with early-onset and stable externalising symptoms (conduct problems and hyperactivity/inattention) were associated with adolescent substance use and antisocial behaviours. Another line of research looks at the role played by internalising symptoms on adolescent risk behaviours. Severe and chronic patterns of antisocial behaviours in adolescence are associated with trajectories of severe and stable internalising problems in adolescent males [42]. A combined externalising and internalising pathway has been found to be associated with more frequent substance use at ages 13 and 14 [43]. In a clinical sample of high-risk adolescents from an inpatient psychiatric unit, internalising symptoms were associated with alcohol use but only when in conjunction with severe externalising symptoms [44]. Accordingly, in our study, a trajectory of combined high externalising and internalising symptoms was associated with higher odds of inclusion in the poly-substance use and antisocial behaviours class. Another study found that internalising symptoms in childhood were negatively associated with adolescent alcohol use [9]. In contrast, a study using cross-sectional data from Nordic countries reported a higher risk of tobacco use in adolescents reporting internalising symptoms [6]. These conflicting results may stem from differences in the onset and stability of internalising symptoms and the varying effects of internalising symptoms on different substances. In our study, a trajectory of high internalising symptoms with preadolescent onset was not associated with any problematic substance use and antisocial behaviours pattern. However, a trajectory of childhood-limited moderate externalising and internalising symptoms was associated with a pattern of alcohol and tobacco use in adolescence.

### Strengths and limitations

We used person-centred statistical methods, specifically growth mixture modelling and latent class analysis. These methods identify latent subgroups within a population to provide an understanding of its characteristic heterogeneity [16]. We modelled trajectories of emotional problems, conduct problems and hyperactivity/inattention jointly. This is better than examining them separately because it allows for a more nuanced patterning of these frequently co-occurring manifestations. Another strength of this study is the use of the large and representative sample from the MCS.

This study is not without limitations, however. We only included mother-reported measures of emotional problems, conduct problems and hyperactivity/inattention and did not account for reported differences between self-report and teacher-report of emotional and behavioural symptoms [45]. We did not account for paternal and peer substance use, which has been shown to be an important influence on adolescent substance use [46]. The trajectories of internalising and externalising symptoms do not represent clinical diagnoses, though the SDQ has shown good predictive power for child and adolescent psychiatric disorders [47]. The higher attrition rates for socioeconomically disadvantaged families and ethnic minorities in the MCS are another source of bias in our study.

### Conclusion

There is a complex interplay between internalising and externalising symptom trajectories and patterns of adolescent substance use and antisocial behaviours. High levels of externalising behaviours and increasing internalising symptoms from early childhood to adolescence are associated with poly-substance use (including alcohol, tobacco and cannabis use) and antisocial behaviour. Additionally, stable and moderate externalising symptoms are associated with substance use and antisocial behaviour in adolescence; this holds true even when these symptoms were limited to childhood. These associations were independent of known risk factors for adolescent substance use and delinquency such as socioeconomic disadvantage and maternal substance use. Interventions on adolescent health promotion and risk behaviour prevention need to address the clustering of substance use and antisocial behaviours as well as the significant influence of early and chronic internalising and externalising symptoms on the aetiology of these behaviours.

## Data Availability

The authors are grateful to the Centre for Longitudinal Studies (CLS), UCL Institute of Education, for the use of these data and to the UK Data Service for making them available. However, neither the CLS nor the UK Data Service bear any responsibility for the analysis or interpretation of these data.

## Acknowledgements

The authors are grateful to the Centre for Longitudinal Studies (CLS), UCL Institute of Education, for the use of these data and to the UK Data Service for making them available. However, neither CLS nor the UK Data Service bear any responsibility for the analysis or interpretation of these data.

